# Nilotinib in Patients with Advanced Parkinson’s Disease: A Randomized Phase 2A Study (NILO-PD)

**DOI:** 10.1101/2020.05.11.20093146

**Authors:** Tanya Simuni, Brian Fiske, Kalpana Merchant, Christopher S. Coffey, Elizabeth Klingner, Chelsea Caspell-Garcia, David-Erick Lafontant, Helen Matthews, Richard K. Wyse, Patrik Brundin, David K. Simon, Michael Schwarzschild, David Weiner, Jaime Adams, Charles Venuto, Ted M. Dawson, Liana Baker, Melissa Kostrzebski, Tina Ward, Gary Rafaloff

## Abstract

**Background:** Nilotinib, a tyrosine kinase Abelson inhibitor, exhibits neuroprotective effects in preclinical Parkinson disease (PD) models.

**Methods:** This Phase 2A double-blind placebo-controlled study in moderate/advanced PD randomized participants 1:1:1 to placebo:150:300 mg nilotinib in matching capsules once daily for 6 months. The primary outcomes were safety and tolerability, the latter defined as ability to complete the study on assigned dose. Secondary outcomes included change in PD disability (Movement Disorder Society Unified Parkinson’s Disease Rating Scale (MDS-UPDRS), Part 3 OFF/ON). Additional exploratory outcomes included serum and cerebrospinal fluid (CSF) pharmacokinetic (PK) profile, and CSF dopamine metabolites.

**Findings:** The study screened 125 and enrolled 76 participants (39% screen failure) between November 2017 and December 2018 at 25 US sites. The last participant completed the study in September 2019. At baseline, mean (standard deviation) age was 64.6 years (7.5), disease duration 9.9 years (4.7), MDS-UPDRS Part 1-3 OFF score 66.4(19.3) and ON score 48.4(16.2), Montreal Cognitive Assessment (MoCA) score 27.1(2.2). Tolerability was 21(84%):19 (76%):20 (77%) in placebo:150:300 mg arm, respectively. Both active doses were safe. The most common reasons for drug suspension were elevations of amylase and/or lipase, which were dose-dependent. The 300 mg group had transitory worsening of MDS-UPDRS-3 ON at 1 month compared to placebo (p<0.01), which resolved by 6 months. There was no difference in the change of MDS-UPDRS-3 OFF from baseline to 6 months between the groups (p=0.17). CSF/serum PK ratio was 0.2-0.3%. There was no evidence of treatment-related elevation of any dopamine metabolites.

**Interpretation:** Both doses of nilotinib were safe and tolerable in these participants, who were selected with strict inclusion/exclusion criteria. There was no evidence of any symptomatic benefit of nilotinib. The drug had low CSF exposure and failed to change dopamine metabolites. These findings do not warrant further testing of nilotinib in PD.

**Funding:** The study was funded by Funded by Michael J Fox Foundation for Parkinson’s Research / The Cure Parkinson Trust / Van Andel Institute. Clinicaltrials.gov NCT03205488

## Introduction

### Background and Objectives

Parkinson’s disease (PD) is the second most common neurodegenerative disease and affects 1% of the population above the age 65(1). Despite numerous prior studies, there are no proven strategies for slowing the PD progression, making this gap a major unmet need(2).

Nilotinib, a BCR-Abelson (c-Abl) tyrosine kinase inhibitor, currently approved for the treatment of chronic myeloid leukemia, has been shown to protect dopamine neurons and prevent accumulation of α-synuclein in animal models of PD(3, 4). Moreover, aberrant activation of c-Abl has been reported in autopsied PD brains and animal models(5). Finally, deletion of the gene encoding c-Abl in mice reduced α-synuclein aggregation and neurobehavioral deficits whereas overexpression of constitutively active c-Abl accelerated α-synuclein aggregation, neuropathology, and neurobehavioral deficits(6). Thus, inhibition of c-Abl has emerged as a therapeutic strategy that has the potential to slow PD progression^(3, 7,8)^. However, based on the intrinsic risk profile of nilotinib and other oncology drugs in this class, safety and tolerability are major factors that will influence the feasibility of development of nilotinib for PD.

At the time of the launch of this study, the only clinical experience with nilotinib in PD was limited to a small, single center open label study(9). That study concluded that nilotinib at tested doses was safe, tolerable, increased dopamine metabolite homovanillic acid (HVA), and decreased phosphorylated c-Abl in the cerebrospinal fluid (CSF) in PD. The study also reported significant symptomatic improvement in motor and cognitive function of the participants. However, the small sample size and open label design of the study precluded firm conclusions, and further investigation was warranted. The primary objective of this study was to assess the safety and tolerability of nilotinib in participants with moderate to advanced PD.

## Methods

### Trial design

This was a 6-month multicenter randomized parallel-group double-blind placebo-controlled trial (clinicaltrials.gov NCT03205488). Potentially suitable participants underwent a screening visit that included detailed clinical assessment, safety laboratory testing and electrocardiogram, and if they continued to qualify, they had a second screening visit to complete a lumbar puncture (LP). Post-screening participants were randomized in 1:1:1 allocation to a once daily dose of either 150mg or 300mg of nilotinib, or matching placebo. Participants and investigators were blinded to treatment assignment. Participants were seen at days 7 and 14 post-randomization and then monthly through the course of the study, with additional safety visits if necessary. Safety was monitored by standard laboratory tests and ECG at all study visits. Final assessment on the study drug was conducted at 6 months after which the study drug was discontinued and participants had safety assessments one and two months after discontinuation of the study drug. Temporary study drug suspensions and re-challenges were allowed based on tolerability and/or pre-specified changes in the laboratory parameters or ECG. Participants who permanently discontinued study drug were terminated from the study. LP was required at screening (prior to randomization) and month 3. An additional LP was optional at one month off the study drug. Detailed trial design and schedule of activities are outlined in the protocol (supplement 1). Recruitment was conducted from November 2017 through December 2018. Last participant completed the study in September 2019. The protocol was approved by the ethics committee at the University of Rochester, which served as the study Clinical Coordination Center, and at each participating site. All participants provided written informed consent. The trial was conducted in accordance with the Principles of the Declaration of Helsinki. Trial monitoring and data management were performed in accordance with the International Conference on Harmonization Good Clinical Practice Guidelines. An Independent Data Safety Monitoring Board (DSMB) reviewed blinded and unblinded data on a regular basis. The authors attest to compliance with the protocol and accuracy and completeness of the data and analyses.

### Setting and Participants

Participants were recruited from 25 Parkinson Study Group (PSG) sites. The PSG is a non-profit consortium of expert Parkinson centers in North America. Participants were eligible to participate in the trial if they had a diagnosis of PD for more than 5 years confirmed by the site investigator based on the established diagnostic criteria(10), were age 40-79, were Hoehn and Yahr stage 2.5 or 3 in the medications ON state(11), were willing to undergo repeated LPs and were on a stable regimen of PD medications that had to include levodopa. Use of monoamine oxidase B (MAO-B) inhibitors initially was exclusionary but was subsequently allowed provided that the dose had been stable for 60 days prior to enrollment. Participants were excluded if they had a diagnosis of atypical parkinsonism, clinically significant depression, history of cardiovascular conditions, liver or pancreatic disease, presence of laboratory or electrocardiographic abnormalities at screening, presence of dementia based on the clinician’s assessment, or a Montreal Cognitive Assessment (MoCA^©^) score < 21 at baseline, or any other conditions or concomitant medications that were associated with increased risk of use of nilotinib as per the drug package insert(12). See full list of inclusion / exclusion criteria in the Protocol.

### Randomization and Interventions

After screening assessments, eligible participants were randomly assigned in a 1:1:1 ratio by a central web-based program to receive a once daily dose of 150mg or 300mg of nilotinib or matching placebo. Nilotinib (150 mg) and matching placebo were provided by the drug manufacturer, Novartis, in kind, and delivered to the University of Rochester Clinical Materials Services Unit which implemented the randomization assignment and delivered study drug kits to the participating sites. Randomization, conducted by the Biostatistics Coordinating Center at the University of Iowa, used random permuted blocks of sizes three and six. All participants started with one capsule daily (150mg or matching placebo) and the dose was escalated to 2 capsules 2 weeks later based on the randomized dose assignment. Participants were instructed to take the study drug at the same time in the morning on an empty stomach as the bioavailability of nilotinib is increased with food, and also to avoid grapefruit products and other foods that are known to inhibit nilotinib metabolism via CYP3A4.

### Outcomes

The primary objective of this study was to assess the safety and tolerability of two doses of nilotinib versus placebo. Tolerability was defined as the percentage of participants who completed the study on their assigned dose across the three study arms. This outcome could be met despite temporary drug interruptions. Conversely, any participant who discontinued study drug, or who completed the study on a dose below their assigned dose, or who failed to complete the study for any reason was deemed not to have tolerated their assigned medication.

Safety was assessed by examining the frequency of treatment-related serious adverse events across all groups within each cohort. Adverse events were collected at every visit through open ended questions and rated by the investigator on severity and causality. An Independent Medical Monitor reviewed all serious adverse events and made adjudications on causality, severity and expectedness.

A key secondary objective was to conduct a futility analysis within each treatment group by comparing the observed change in the Movement Disorder Society Unified Parkinson’s Disease Rating Scale (MDS-UPDRS)(13) Part III “ON” score between baseline and month 6. Other secondary objectives of the study were to 1) establish the degree of symptomatic effect of nilotinib as measured by change in MDS-UPDRS part III (motor) in the PD medications ON state (approximately one hour after a dose of PD medication) between baseline and Visit 2 (1 month) and final visit on study drug and 30 days off study drug; 2) To explore the impact of nilotinib on progression of PD disability as measured by the change in the MDS-UPDRS Part III score in the defined medications OFF state (at least 12 hours after the last dose of PD medications) between baseline and 6 months. A number of clinical exploratory outcomes measuring change in disability, quality of life and functional status from baseline to 6 months were pre-specified in the statistical analysis plan (SAP), see supplement 2.

### Exploratory pharmacokinetics and biomarkers samples and analysis

During study visit month 3, pre-dose trough and post-dose serum samples were collected for pharmacokinetic assessment at steady-state. The post-dose serum sample was collected approximately 2 hours following administration of an in-clinic dose to approximate the time at which maximum concentration (Cmax) is achieved in the serum (reported T_max_ ~2-3 hours(12, 14). A corresponding CSF sample was also collected 2 hours post-dose. Total serum and CSF concentrations of nilotinib were measured using validated liquid chromatography with tandem mass spectrometry assays (WuXi AppTec, Shanghai, China). The lower limits of quantitation for the serum and CSF assays were 2.50 and 0.200 ng/mL, respectively. Additional serum samples were collected at other study visits for future pharmacokinetic analysis (see Protocol). Serum and CSF samples were analyzed to: (a) determine the serum PK and exposures of nilotinib at steady state, (b) investigate the ability of nilotinib to cross the blood brain barrier and (c) correlate CSF levels with the clinical and biomarker outcomes.

For the biomarkers, we assayed phospho-cAbl levels in the CSF using PathScan® pan-tyrosine ELISA, as reported by Pagan et al^9^. Levels of dopamine, its metabolites, other monoamine neurotransmitters (norepinephrine, epinephrine, serotonin, histamine) and their metabolites, were tested using an analytically validated mass spectrometric multiplexed assay of 17 analytes (Supplement 3). Nilotinib-induced changes in each biomarker were determined by comparing values at the screening visit versus those at 3 months, as well as by comparing the values in the placebo group with each nilotinib arm. Additional blood, serum, plasma, DNA and CSF samples were stored for future exploratory research. See supplement 4 for detailed summaries of the biologic samples collection and processing.

### Statistical Methods

The first objective was assessed by comparing the proportion of study participants who met the study definition of tolerability among the placebo group and each of the treatment groups, with a one-sided Fisher’s exact test using a significance level of 0.05 used for each test.

The safety objective was primarily assessed by comparing the proportion of study participants with any treatment-related SAE among the placebo group and each of the treatment groups using a Fisher’s exact test. To provide an overall summary of the safety of each nilotinib dose, additional safety assessment involved further comparisons of AEs and SAEs across the three groups. The safety and tolerability information were synthesized and reviewed by the steering committee at the end of the study.

The key secondary objective involved a single group hypothesis within each PD group to assess “futility” for replicating the large difference observed in the prior study(9). That study observed a mean change over 6 months in the 150 mg nilotinib group of 7.0 (SD = 12.9) points on the UPDRS(15). Since we used the MDS-UPDRS, we included a correction factor of 1.4 as specified in Goetz et al(16). Thus, this objective was assessed by conducting a futility test within each nilotinib dose group based on a null hypothesis that the observed difference was less than a 9.8 unit reduction on the MDS-UPDRS (7.0 x 1.4) versus an alternative that the reduction was less than 9.8 units (or an increase was observed over time). Rejecting the null hypothesis would imply that a future study is unlikely to observe the large reductions previously reported. However, that would not rule out potentially meaningful effects on a smaller scale. This null hypothesis was evaluated based on an assessment of parameter estimates from a non-linear mixed effects model of change from baseline with time a categorical variable and baseline MDS-UPDRS and calculated levodopa equivalent daily dose (LEDD) included as covariates(17).

Similar linear mixed models were used to compare other secondary and exploratory objectives involving the MDS-UPDRS. For each comparison, a global two degree of freedom test was initially used to test for any differences among the three groups. If the global test was significant, then stepdown pairwise comparisons were used to further explore any observed differences. Similar non-linear mixed models were used to assess the exploratory PDQ-39 and LEDD outcomes. The categories for the Clinical Global Impression Scale (CGI) score for both the participant and investigator were collapsed into three categories (improved, stayed the same, worsened) and were compared using a Fisher’s exact test. Pharmacokinetic and biomarker results were summarized by nilotinib dose level using geometric means and 95% confidence intervals (CI). Association between CSF nilotinib concentrations and dopamine turnover indices were assessed by Spearman correlation.

### Sample Size

We assumed that at least 90% of participants on placebo would meet the study definition of tolerability, and that an absolute decrease of 30% or greater with respect to tolerability for active treatment arms versus placebo would provide sufficient tolerability concerns that would not warrant further study of that dose. Hence, the sample size was chosen to provide sufficient power to compare an expected 90% vs. 60% or lower tolerability rate in the placebo group versus each of the treatment groups. Under these assumptions, a total of 75 participants provided at least 80% power at the one-sided 0.05 significance level.

#### Role of the funding agency

This study was funded by the Michael J Fox Foundation for Parkinson’s Research (MJFF), The Cure Parkinson Trust and Van Andel Institute. Research officers (BF, RW, PB, HM) from the three funding agencies were involved in the study design, interpretation of results, review/revision of this manuscript, and decision to submit this manuscript for publication and are listed amongst the authors. Novartis provided the drug and placebo supply as in kind contribution but were not involved in the study design or execution.

## Results

### Participants

Of 125 participants screened for eligibility, 76 were enrolled and randomized: 25 to nilotinib 150mg: 26 to nilotinib 300mg and 25 to placebo. There was a high percent of screen failures (49/125) (Figure 1). The major reasons for exclusion were comorbid conditions (12), exclusionary ECG (10), cardiovascular conditions (7) and low HY stage (7). Baseline characteristics of the enrolled cohort are detailed in Table 1. The groups were generally well balanced, but disease duration was slightly higher in the nilotinib 300mg group and age lower in nilotinib 150mg group (Table 1). There were 8 premature withdrawals: one in the placebo arm (tremor), 2 in the nilotinib 150mg arm (anxiety and increase in lipase) and 5 in the nilotinib 300mg arm [increase in lipase (2), arthritis, arrhythmia and an abnormal ECG]. The latter was present at screening but was not identified until the participant was randomized so was not considered treatment related.

**Table 1:**
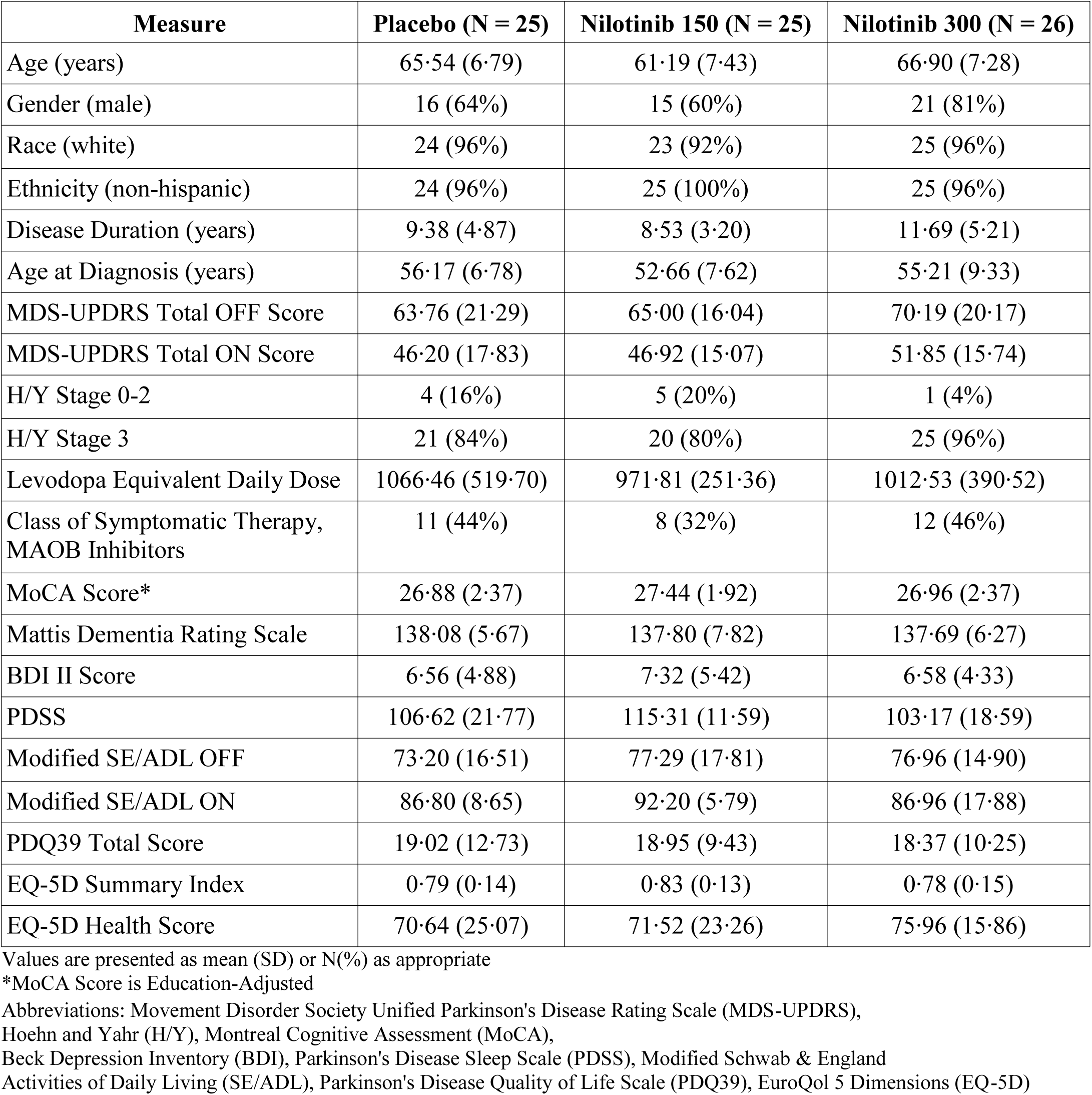
Baseline Demographics and Disease Characteristics.

**Figure 1.**
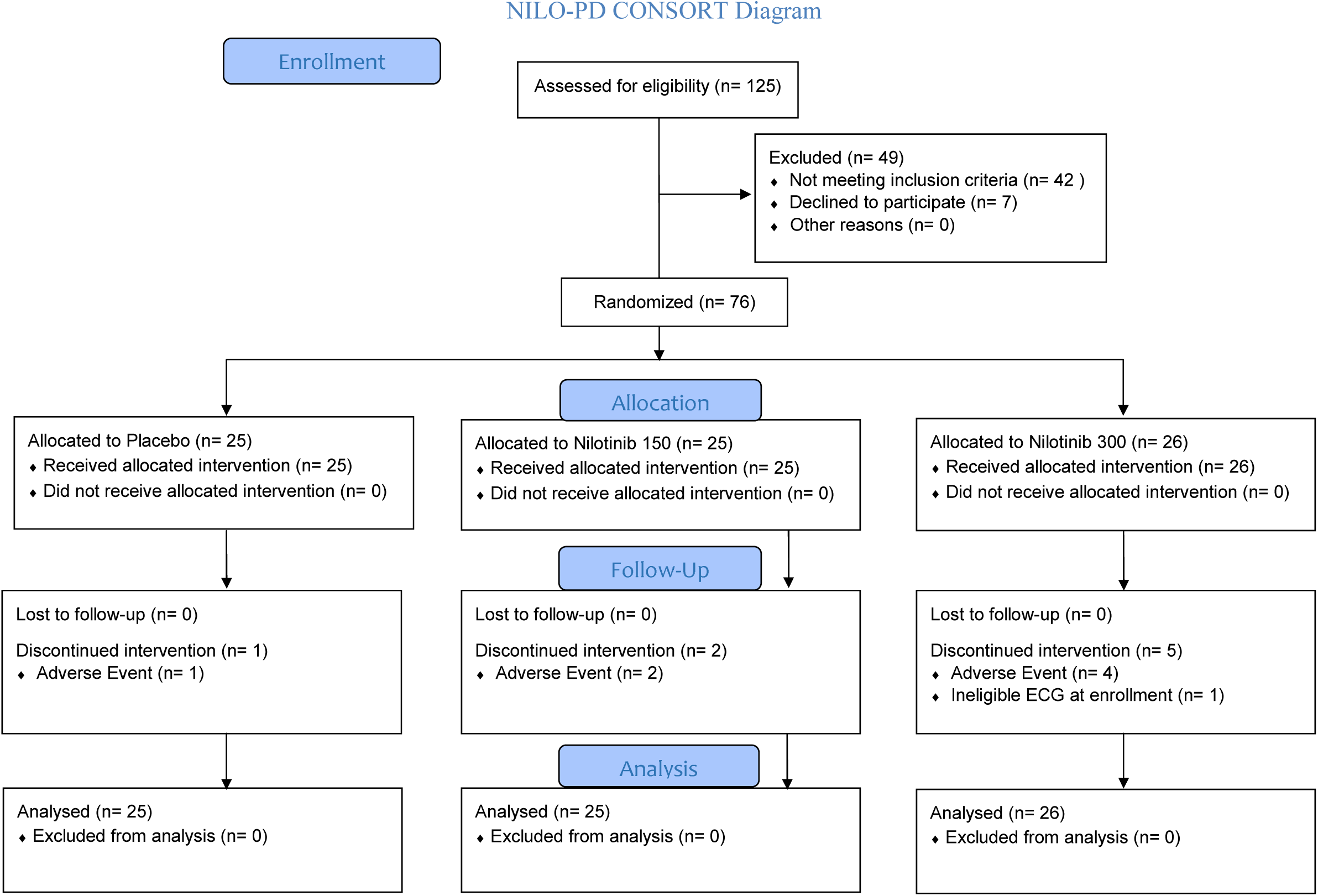

Both doses of nilotinib were tolerable, with 21(84%): 19(76%): 20(77%) meeting the study definition of tolerability for the placebo: nilotinib 150 mg: nilotinib 300 mg, respectively (p=0.36 for nilotinib 150mg and p=0.39 for nilotinib 300 mg, respectively, versus placebo; Table 2). However, there were more premature withdrawals due to adverse events in the nilotinib 300mg group (Figure 1). The most common reasons for dose reduction or temporary suspension were increase in lipase and /or amylase with no significant imbalance between the groups and no associated clinical symptoms. There was no difference in the numbers of serious adverse events between the groups: 2 in placebo: 1 in nilotinib 150mg: 1 in nilotinib 300mg groups. Only one serious adverse event was considered treatment related (arrhythmia in nilotinib 300mg group). Adverse events presented in the order of frequency are listed in Table 2.

**Table 2:**
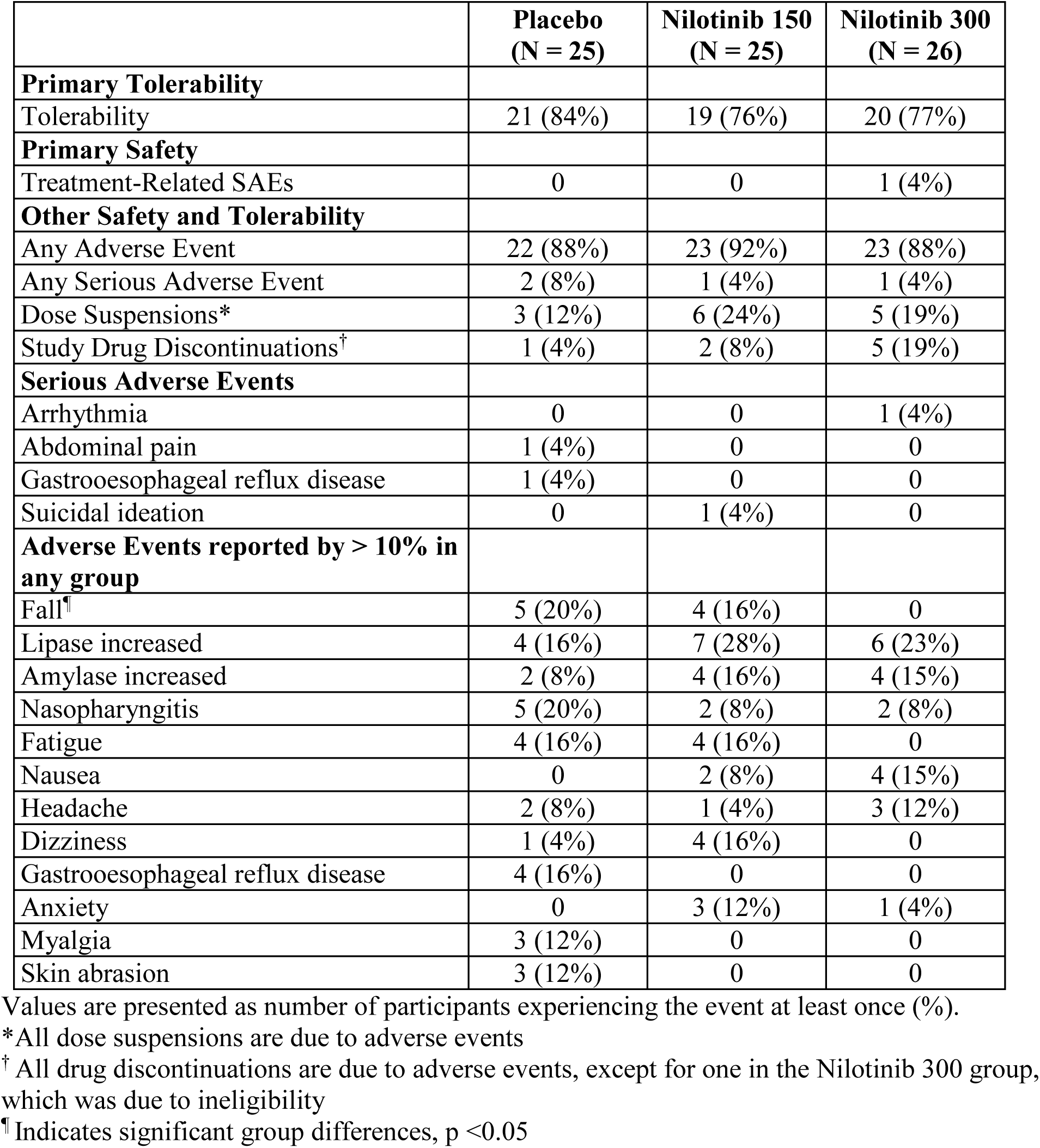
Safety and Tolerability Outcomes.

### Secondary outcomes

Change of MDS-UPDRS Part III (motor) in the medications ON and OFF state are presented in Table 3 and Figure 2. Both analyses demonstrate “futility” from the previously observed large symptomatic effect in Pagan et al(9) (p < 0.001 for both comparisons), suggesting that a future study is unlikely to observe such a large symptomatic effect. There were no significant differences observed between the groups in the medications ON state from baseline to month 6 (p=0.0768), and month 6 and 1 month off the study drug (p=0.4657). Interestingly, a significant difference was observed across groups from baseline to month 1 (p = 0.0306). Stepdown pairwise comparisons suggest that this was primarily driven by a transitory worsening in nilotinib 300mg group compared to placebo at month 1 (p = 0.008). There was no significant difference observed between the groups in the change of MDS-UPDRS Part III in the medications OFF state between baseline and month 6 (p=0.1711). No significant differences were observed for any of the exploratory clinical outcomes (Table 3).

**Table 3:**
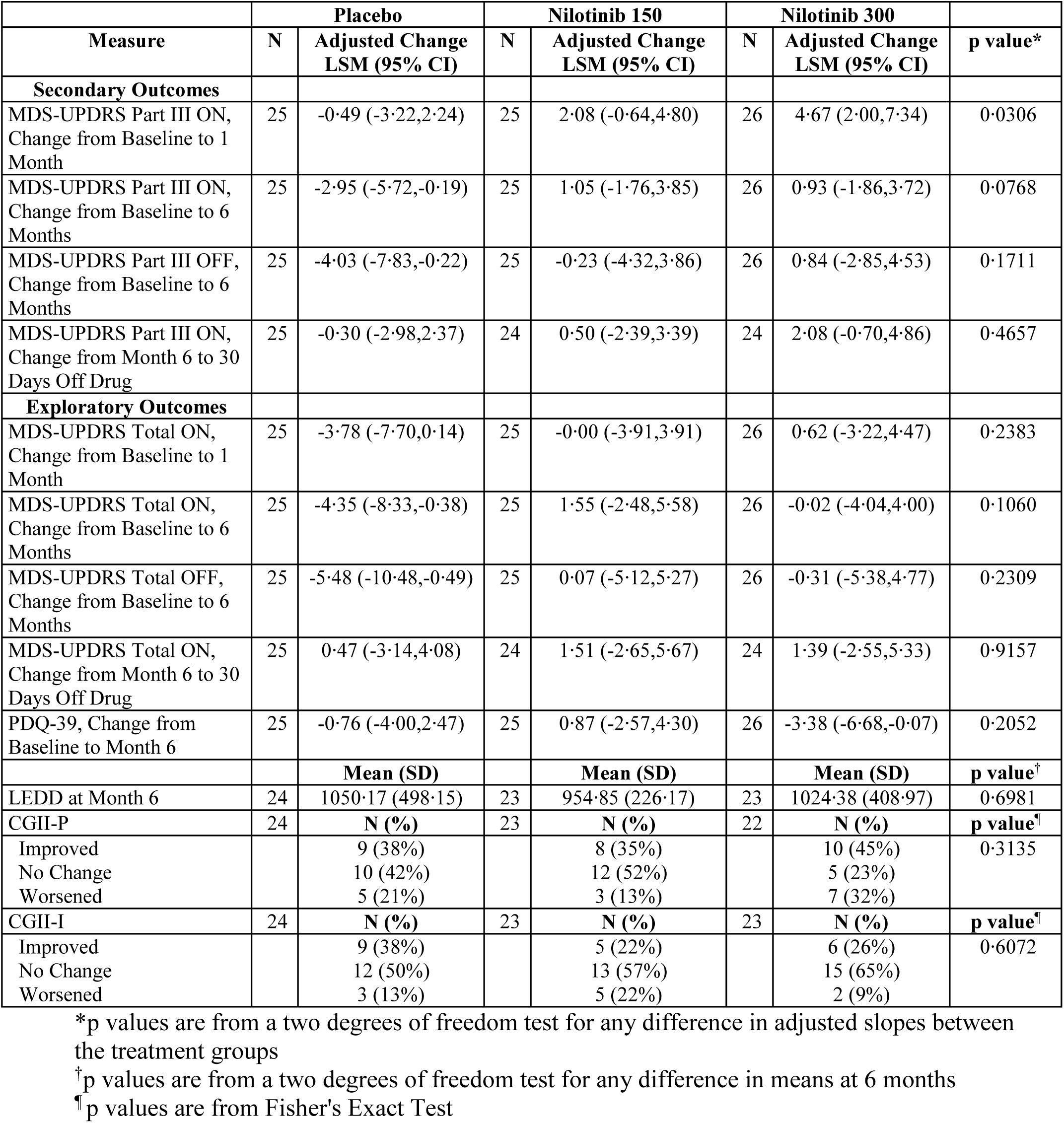
Changes in Secondary and Exploratory Outcomes Over Time.

**Figure 2:**
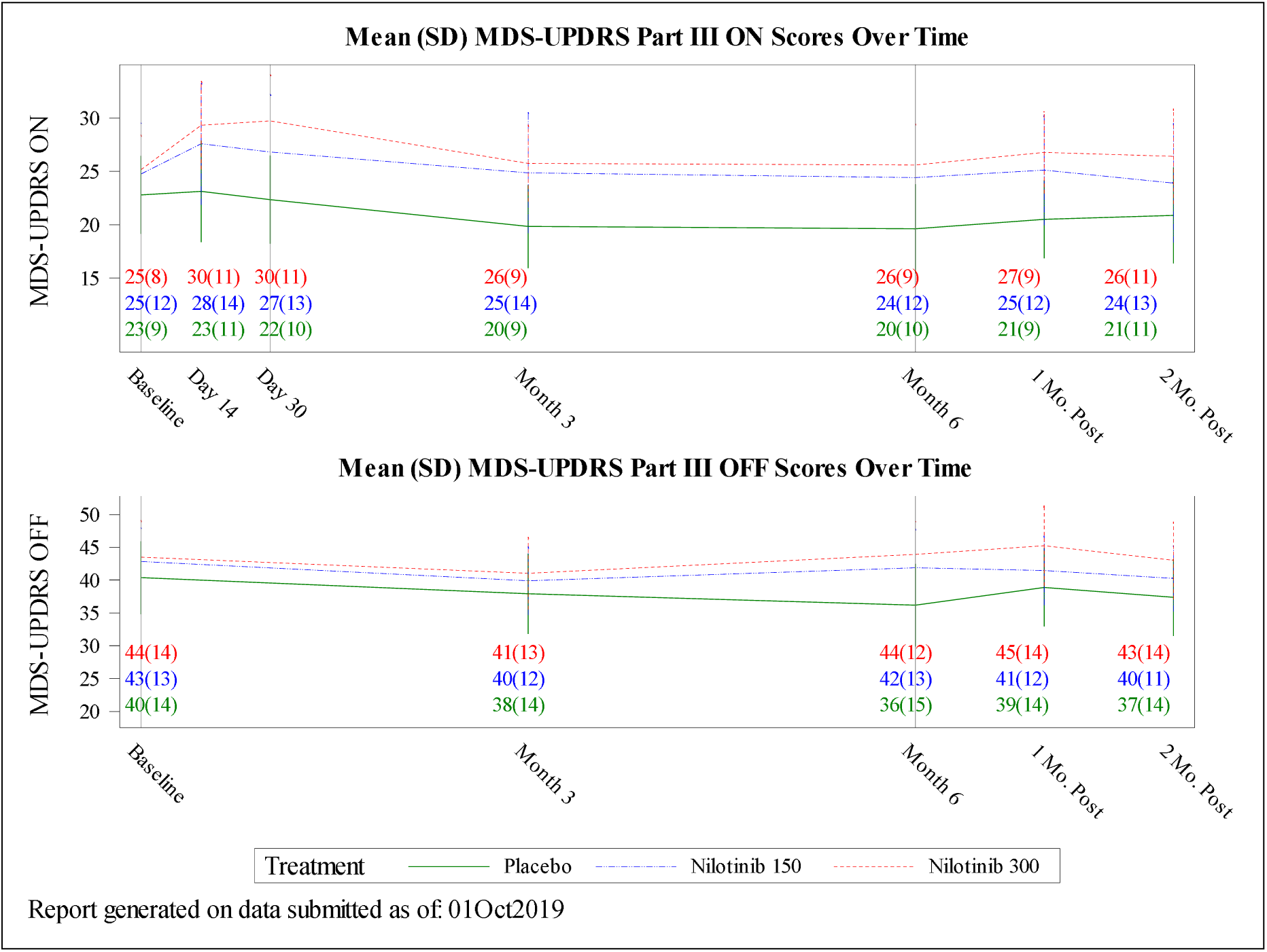
Movement Disorder Society Unified Parkinson’s Disease Rating Scale (MDS-UPDRS) Part III ON & OFF Over Time.

### Exploratory pharmacokinetic and pharmacodynamics outcomes

Nilotinib serum and CSF concentrations at month 3 are presented in Table 4. Total serum concentrations were within range of the previously reported values from nilotinib investigator brochure(12). Concentrations of nilotinib in the CSF collected at 2+/-0.5 hours (~T_max_) post dose were 0.19% and 0.26% of that in the serum for the 150mg and 300mg doses, respectively. The absolute concentrations observed at ~T_max_ were only 8-13% of the reported cellular half-maximal inhibitory concentration (IC_50_) of 20 nM (11 ng/mL) for inhibition of c-Abl by nilotinib^(18)^.

**Table 4:**
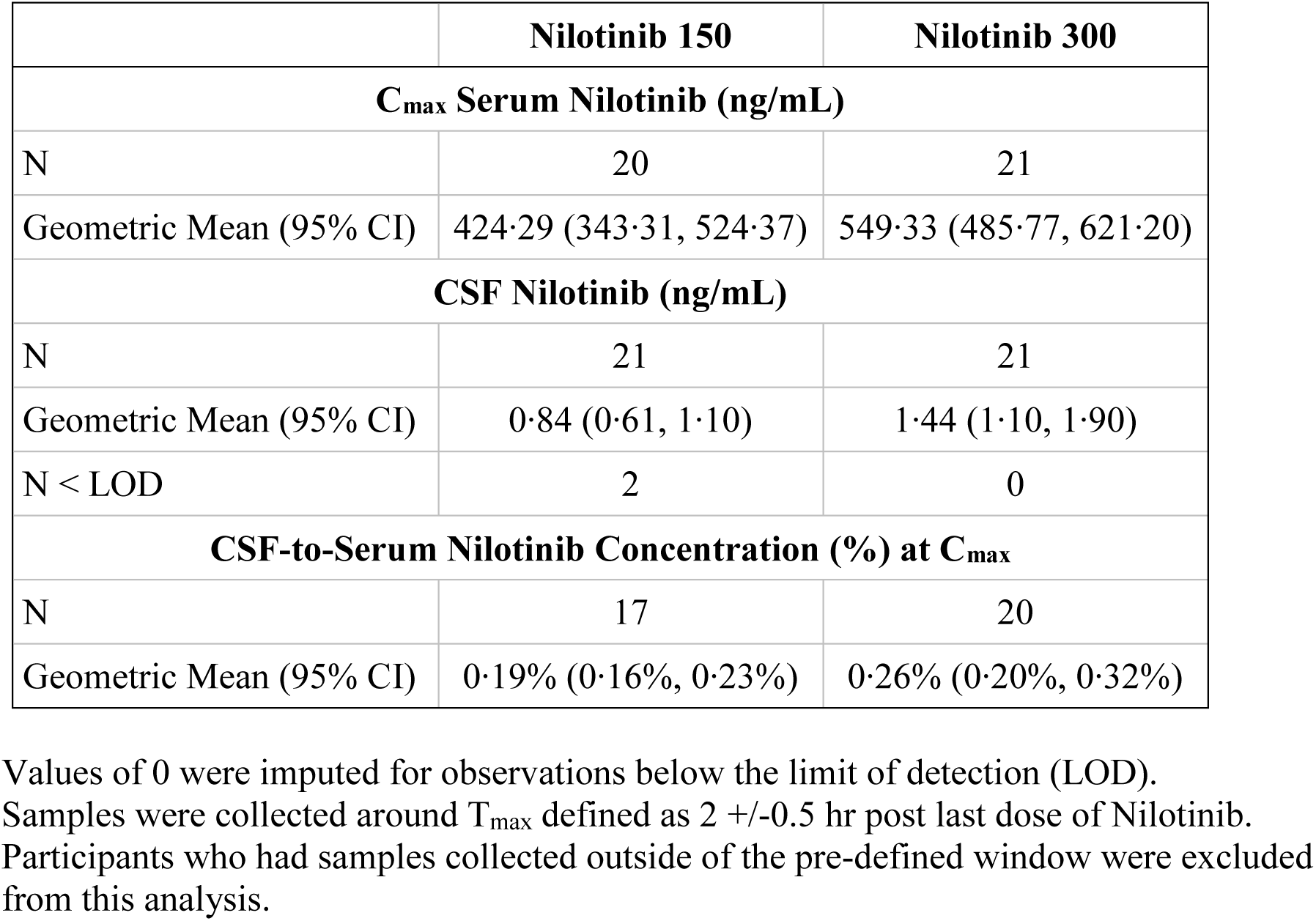
Steady State Nilotinib Serum and CSF Concentrations at 3 Mon around T_max_ (2 +/-0.5 hr)

We tested CSF phospho-c-Abl using methods reported by Pagan et al(9) in a pilot study of 20 CSF samples from healthy controls and PD donors and could not detect the analyte in any biospecimen leading to the decision of not measuring this analyte in CSF samples from the present study (data not included).

Three-month nilotinib treatment did not alter CSF levels of any monoamine or its metabolite measured in samples collected at the expected T_max_ of 2 h +/-0.5 h post-dose (Figure 3, panel A-C). In order to ensure that concurrent treatment with MAO-B inhibitors did not affect the biomarkers examined, we repeated the analyses after excluding MAO-B treated participants. These analyses also failed to show changes in dopamine or its metabolite levels after nilotinib treatment (Supplementary Table 1). Furthermore, there was no correlation between the CSF nilotinib levels and levels of dopamine metabolites or their ratios with dopamine (Figure 3, Panel D and E).

**Figure 3:**
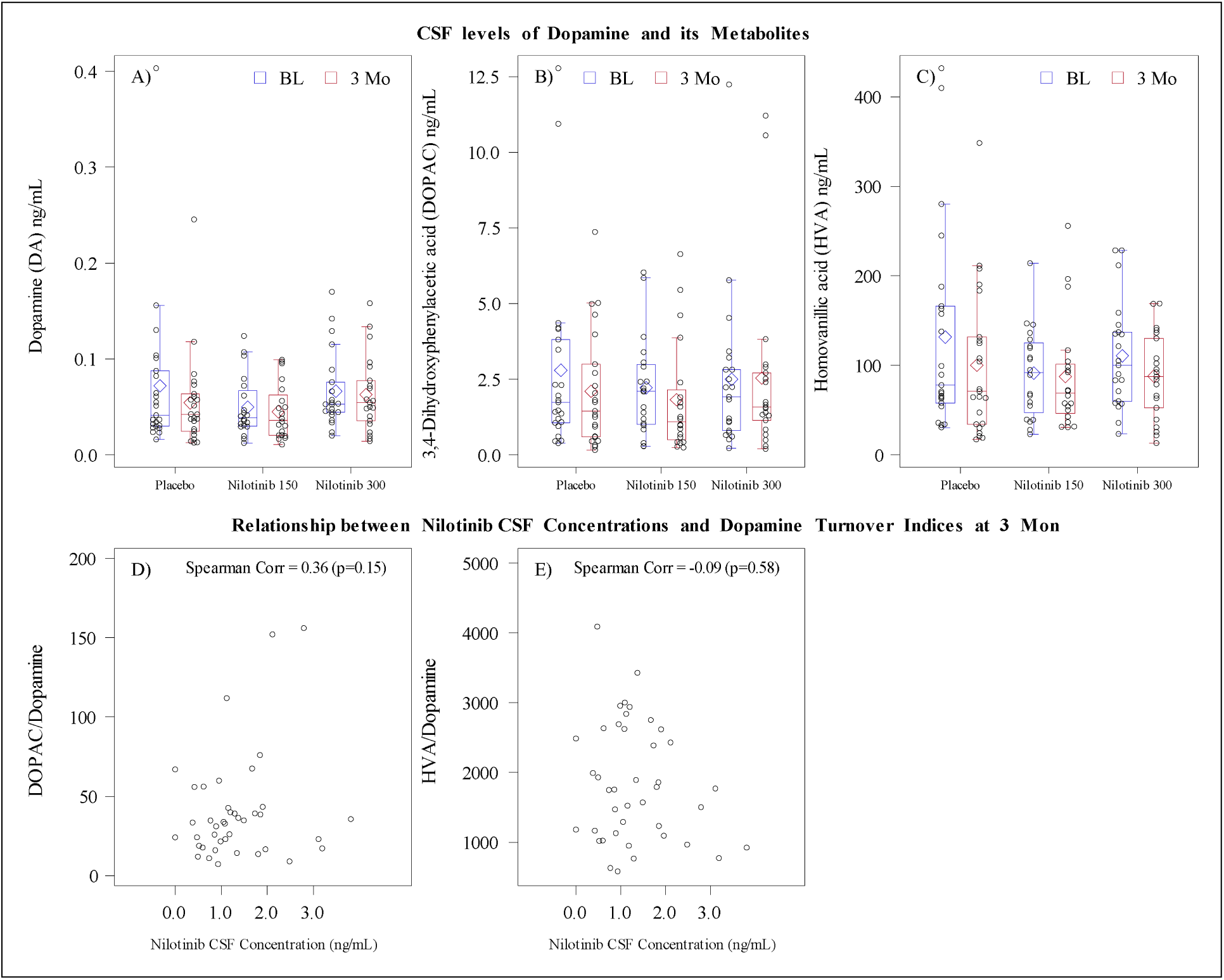
CSF levels of Dopamine, its Metabolites & the Relationship between Nilotinib CSF Concentrations & Dopamine Turnover Indices.

In parallel to the human study, we conducted a study in chronically cannulated beagle dogs to assess whether at steady state nilotinib crosses the blood-brain barrier sufficiently and inhibits c-Abl in the brain (Supplement 5). The absolute levels of nilotinib in the CSF at the two doses tested (20 mg/kg and 50 mg/kg per day for 2 weeks) were ~8-10 times higher than those observed in our clinical study. Consistent with the human data, the dog study demonstrated CSF exposures that were <1% of those seen in the serum. The brain levels of unbound nilotinib were 2-4% of serum levels in the dog and fell below the cellular IC_50_ for c-Abl inhibition. In accordance with the apparent poor brain penetration, we failed to detect a decrease in p-Abl levels in the brain as measured by immunoblots.

## Discussion

Our study demonstrates acceptable safety and tolerability of both tested doses of nilotinib in participants with moderately advanced PD. This is an important conclusion as nilotinib has a significant risk profile including a black box warning for the risk of cardiac arrhythmias (QT prolongation) and sudden death as well as a number of safety concerns regarding risk of myelosuppression, cardiovascular occlusive disease, electrolyte abnormalities, hepatotoxicity, pancreatitis and others. While it is reassuring that we did not observe significant increase in serious or overall adverse events in nilotinib treated groups, it also has to be acknowledged that study participants were selected based on stringent screening that excluded participants with history or evidence of any conditions that would increase the risk of their exposure to nilotinib. As such the study had a 39% screen failure rate, higher than the projected and usually observed 25% rate in other PD studies. In addition, participants had stringent laboratory and ECG monitoring through the course of the study which contributed to the participant burden. We did not observe any significant hematological adverse events that would have been expected based on the mechanism of action of the drug, likely related to the fact that we used lower doses than indicated for the drug’s primary indication.

In regard to the secondary efficacy analysis, we did not observe any significant impact of either dose of nilotinib on PD disability. The rationale for focusing on symptomatic effect of nilotinib in this early phase safety study was based on a significant symptomatic effect on motor and cognitive function reported in the prior open label study(9). We also did not observe trends for symptomatic benefit of either dose of nilotinib on overall PD disability (MDS-UPDRS total score) OFF and ON, severity of motor complications (MDS-UPDRS Part IV), cognition, sleep, or quality of life. Our results are in line with the conclusions regarding safety, tolerability and lack of the symptomatic effect of nilotinib from another recently completed study that tested the same doses in a similar population but extended the duration of follow up to 12 months(19). Results of both studies highlight limitations of any read out of the symptomatic effects in the open label studies. Lack of a short-term symptomatic effect is not surprising as the mechanism of action of nilotinib as a putative PD disease modifying intervention is linked to improved cell autophagy and, if effective, would be expected to change the trajectory of the disease progression rather than have an immediate benefit. This study was not designed or powered to address long term benefits of nilotinib. We had pre specified “go”/”no-go” criteria to inform whether this study results were sufficient to move forward with the testing of nilotinib in a de novo PD cohort. The results from the primary, secondary, and exploratory analyses reported here informed a steering committee decision not to move forward.

Assessment of the pharmacokinetic profile and impact of nilotinib on the biomarkers panel, while exploratory, is extremely important in interpreting of the study results. While nilotinib serum PK data were previously published(14), there were no published data on CSF penetration of the drug in PD patients at steady state, aside from the small cohort from the open label study^9^. Defining CNS penetration was essential for the decision regarding further development of nilotinib for PD indication. Our data demonstrate serum concentrations that are in line with the previously reported data but very low CSF concentrations, with 0.19-0.26% CSF to serum ratio. The CSF concentrations are about 10 times below the IC_50_ for c-Abl inhibition in cell assays reported to be at 20 nM(18). IC_50_ is a measure of potency of a drug in inhibiting its specific biological target. The observed CSF concentrations of nilotinib indicate that minimal c-ABL inhibition would be achieved at doses within the safe therapeutic range. We attempted direct testing of c-Abl inhibition but there are no validated assays.

As a measure of downstream effects of nilotinib reflecting improvement of dopaminergic cell function we measured a panel of dopamine, dopamine metabolites and other monoamines. We did not observe a change in any of these biomarkers as measured at the 3-month time point. We also did not see a correlation between CSF nilotinib exposure and dopamine or dopamine metabolites levels. These results indicate that nilotinib does not alter these indices of dopaminergic cell function, potentially because it does not achieve a sufficient concentration in the CNS. Dopamine metabolites are sensitive to the effect of PD dopaminergic therapy(20). Specifically, treatment with MAO-B inhibitors can reduce the level of homovanillic acid (HVA), the major dopamine metabolite, and potentially impact interpretation of the results. Our analysis controlled for the dose and change in the dopaminergic therapy. Considering that about 30% of participants in our study were receiving MAO-B inhibitors, we ran a post-hoc subgroup analysis excluding these participants. That analysis did not change the conclusions. Our analysis of dopamine metabolites differ from the ones reported by Pagan et al(9, 19, 21) where they found an increase in CSF HVA level in 150mg but not in 300mg dose group. These differences might be attributed to the methodological differences.

Our biomarkers panel did not include measurement of α–synuclein or tau. While appropriate CSF samples were collected and stored, the low CNS penetration of nilotinib suggested that the plausibility of observing an impact of the drug on these biomarkers was low.

### Limitations

The study has a number of limitations. The study was conducted in participants with moderately advanced PD which is not the population typically targeted for PD disease-modifying interventions. The rationale for selecting that profile of PD was based on the need to test the large symptomatic effect seen in the previously reported study by Pagan et al(9) as well as to test safety and tolerability of this drug in a more vulnerable PD population. Our study had 6-months duration of intervention compared to 12 months in the recently reported study by Pagan at el(19). Considering that the primary study objectives were to determine safety and tolerability, we believe that 6 months exposure was sufficient to address these questions as well as our secondary objectives to assess degree of symptomatic effect of nilotinib. The study was not designed to address the question of a disease modifying effect of nilotinib. The dose of nilotinib was selected in the lower range of approved doses and, as such, was expected to have lower serum and corresponding CSF concentrations but, based on the safety profile of nilotinib, we do not expect that testing higher doses will be feasible.

In conclusion, while we demonstrated acceptable safety and tolerability of nilotinib in the carefully selected cohort of participants with moderately advanced PD, low CSF exposure of the drug and failure to impact levels of dopamine and its metabolites, we concluded that nilotinib is not suitable for further testing in PD. These results do not refute the hypothesis that c-Abl inhibition is a potential important therapeutic target for PD neuroprotective interventions. Indeed, there are a number of novel molecules targeting c-Abl pathway in development that have a better therapeutic profile. Results of these studies are highly anticipated.

**Supplementary Table 1.**
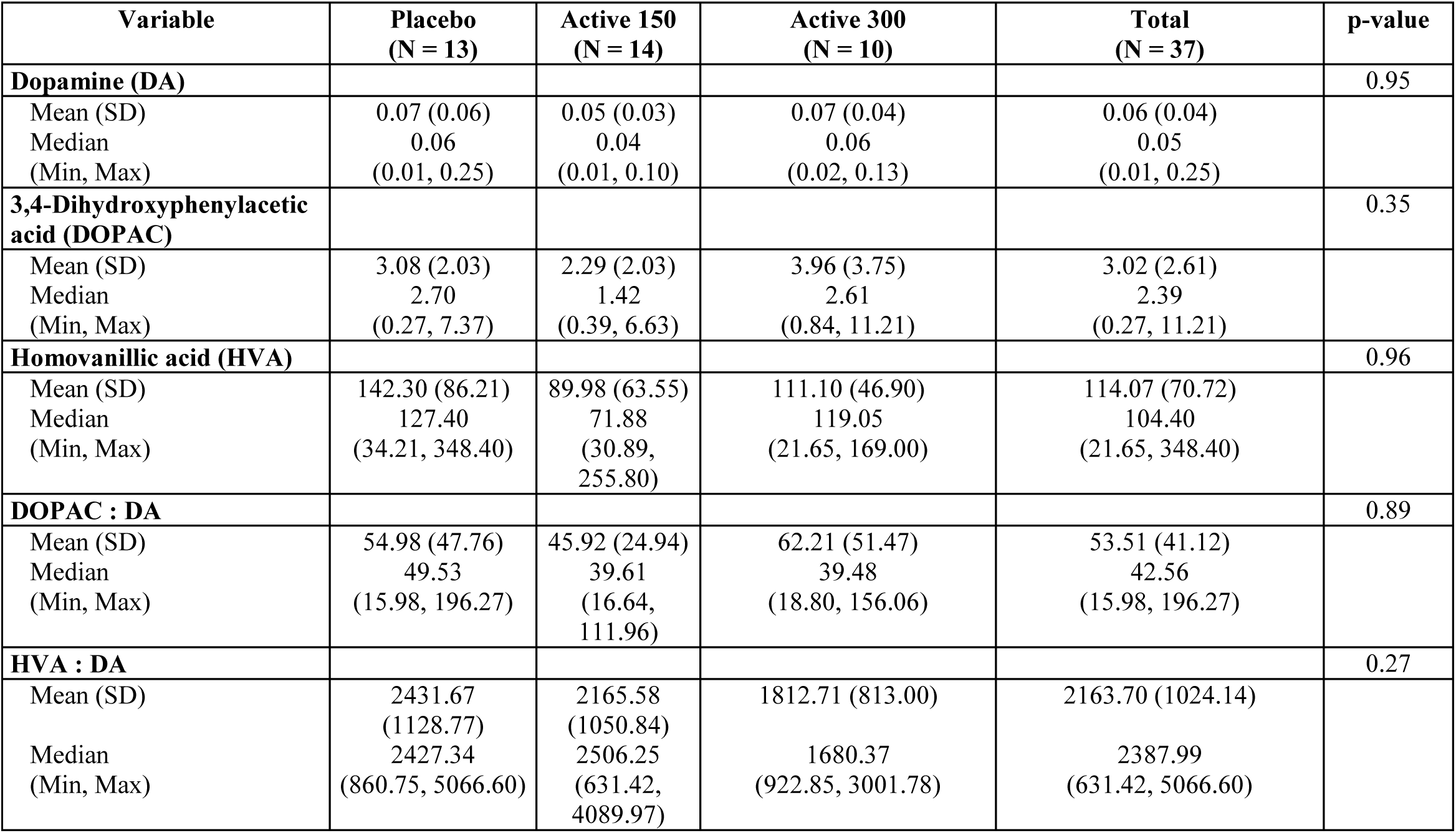
CSF levels of Dopamine, its Metabolites by Treatment Group Excluding Participants on MAO-B Inhibitors.

## Data Availability

Available

## Supplementary Materials

Supplement 1. Protocol

Supplement 2. Statistical analysis plan

Supplement 3. Analytical method for assessment of monoamines and their metabolites in the CSF.

Supplement 4. Analytical methods to determine nilotinib concentrations in the serum and CSF.

Supplement 5. Serum and CSF Pharmacokinetic Study in the Dog.

Supplement 6. List of Non-Author contributors (to be listed so they can be searched in PubMed).

### Supplement 3. Analytical method for assessment of monoamines and their metabolites in the CSF

The CSF biospecimens were analyzed by Biocrates Life Sciences AG (Innsbruck, Austria) using a validated multiplexed, mass spectrometry-based analytical platform. The table below shows the 17 distinct analytes measured and their respective Lower Limit of Detection (LOD) and Upper Limit of Quantification (ULLQ) values. Absolute concentrations of monoamines and related metabolites were determined using procedures adapted from Yamada et al(22). In brief, samples were subjected to ultracentrifugation and derivatization prior to online solid-phase extraction and LC-MS/MS analysis (Symbiosis Pharma, Spark, Emmen, Netherlands) coupled to an Applied Biosystems API4000 MS/MS-System). Heavy isotope-labeled catecholamines used as internal standards were added to the standard calibration curve as well as to each sample before extraction to correct for random and systematic errors. Each analyte was normalized to its appropriate internal standard, resulting in relative areas.

|  | Metabolite Name | LOD [ng/mL] | ULOQ [ng/mL] |
| --- | --- | --- | --- |
| Catecholamine catabolites | 3,4-Dihydroxymandelic acid | 0.3 | 25 |
|  | 3,4-Dihydroxyphenylacetic acid (DOPAC) | 0.05 | 100 |
|  | 3,4-Dihydroxyphenylglycol (DOPEG) | 0.01 | 40 |
|  | 3-Methoxytyrosine | 0.4 | 300 |
|  | 3-O-Methyldopamine | 0.02 | 7.5 |
|  | 4-Hydroxy-3-methoxymandelic acid | 0.3 | 400 |
|  | 4-Hydroxy-3-methoxyphenylglycol (HMPG) | 0.03 | 150 |
|  | Homovanillic Acid (HVA) | 0.5 | 500 |
|  | Metanephrine | 0.015 | 7.5 |
|  | Normetanephrine | 0.02 | 35 |
| Catecholamine pre-cursor | 3,4-Dihydroxyphenylalanine (DOPA) | 0.05 | 19.3 |
| Catecholamines | Adrenaline (Epinephrine) | 0.01 | 2.5 |
|  | Dopamine | 0.005 | 1.8 |
|  | Noradrenaline (Norepinephrine) | 0.04 | 10 |
| Signal Substance, Serotonin & Metabolite | Histamine | 0.05 | 200 |
|  | 5-Hydroxy-3-indoleacetic acid (5-HIAA) | 0.5 | 128 |
|  | Serotonin (5-HT) | 0.1 | 87.5 |

### Supplement 4. Analytical methods to determine nilotinib concentrations in the serum and CSF

Bioanalytics for nilotinib concentrations in serum and CSF was performed at WuXi AppTec (Shanghai, China) under Good Laboratory Practice (GLP) procedures. Frozen biospecimens aliquots were shipped to WuXi on dry ice and stored at −80°C until analysis of nilotinib concentrations by previously validated LC-MS/MS assays for each matrix. Concentrations of nilotinib were determined using the slope and intercept of calibration curves for each matrix as described below. In addition, quality control samples were included in each batch analyses and data were accepted only if calculated concentrations of the quality control samples were within 15% of their nominal values. The lack of carryover was demonstrated by injecting blank samples after nilotinib injected at Upper Limit of Quantification for each matrix. Incurred sample reproducibility was conducted for each matrix using 10% of study samples to demonstrate the reproducibility of nilotinib concentration in the incurred samples under study storage conditions.

#### Serum Assay

The reference material was nilotinib and internal standard was [^13^C3^15^N_2_] AMN107, supplied by Novartis. Human serum was used as a blank matrix for preparation of 8-point calibration curve (2.5 to 5000 ng/mL nilotinib). The Lower Limit of Quantification (LLOQ) and Upper Limit of Quantification (ULOQ) were 2.5 ng/mL and 5000 ng/mL, respectively. Freshly prepared quality control samples (6, 150, 1400 and 4000 ng/mL nilotinib) were tested in each batch.

#### CSF assay

The reference material was nilotinib and internal standard was nilotinib-d6. Triton X-100 (20 mg/mL) was added to artificial CSF (Harvard Apparatus, Cambridge, MA, USA) to reduce non-specific binding to tubes and used as blank matrix. An 8-point calibration curve (0.200 to 100 ng/mL) was prepared in the blank matrix. The LLOQ and ULOQ for CSF were 0.200 ng/mL and 100 ng/mL, respectively. The quality control samples (0.6, 4, 40 and 75 ng/mL nilotinib) were prepared fresh in human CSF with Triton X-100 (20 mg/mL) and were tested for each batch of CSF analyzed.

#### Pharmacokinetic Sampling Study Design

On day 14 and months 1 and 2 of the study, pre-dose trough serum samples were collected before administration of the daily dose of study drug. Serum samples were also collected at 2 hours post dose (reported as T_max_) at month 3 to approximate a maximum concentration (C_max_), and serum samples at random times relative to dose at months 2, 4, and 6. Serum was also collected in month 7 after a month off the study drug. CSF samples were collected at screening (i.e., pre-drug) 2 hours post dose of study drug at month 3 and a month off the study drug (month 7); the latter CSF collection was optional.

### Supplement 5. Serum and CSF Pharmacokinetics (PK) Study in the Dog

The objective of this study was to establish steady-state PK of nilotinib in the serum and CSF as well as brain exposure at ~Tmax to evaluate its penetration into the CNS.

### Methods

#### Animals and Surgery

The study was conducted at MPI Research (Mattawan, MI, USA) under a protocol approved by the Institutional Animal Care and Use Committee (IACUC). Animal welfare was in compliance with the U.S. Department of Agriculture’s (USDA) Animal Welfare Act (9 Code of Federal Regulations (CFR) Parts 1, 2 and 3), the Guide for the Care and Use of Laboratory Animals, Institute of Laboratory Animal Resources (National Academy Press, Washington, D.C.). Ten adult male beagle dogs weighing 9-12 Kg were surgically instrumented with a jugular vein catheter for blood collection as well as an intrathecal catheter between L3 and L4 with an access port for collection of cerebral spinal fluid (CSF) under general anesthesia.

#### Nilotinib treatment

Dogs were randomly assigned to 2 treatment arms of 5 animals each and received either 20 mg/kg or 50 mg/kg nilotinib in a formulation of 1.5% Avicel/0.3% HPMC in water *via* oral gavage daily for 14 days (dose volume = 5 mL/kg). The lower dose was selected on the basis of dog PK data (Novartis Investigators’ Brochure) to target serum PK in the range of human PK at 300 mg of nilotinib. The animals were not fasted prior to dosing and food/fluids were provided *ad libitum*.

#### Biospecimen Collection and Bioanalytics

Blood and CSF samples were collected from 4 animals of each dose group on Day 14. Blood collection from the jugular vein occurred at pre-dose, and 0.25, 0.5, 1, 2, 4, 8, 12, 24 and 48 hours post-dose and was processed for serum collection. CSF collection occurred at pre-dose, and 1, 2, 4 and 12 hours post-dose. Dosing of the animals was continued on day 15 and 16 as per their originally assigned dosage. On day 16, animals were euthanized 2 hours post-dose (~Tmax) for collection of brain tissue. An additional 2 treatment-naïve dogs without catheterization were necropsied as controls for the brain exposures. All biospecimens were frozen and stored at −60 to −90°C until analyses. Nilotinib concentrations in serum, CSF and brain homogenate were assessed by liquid chromatography-tandem mass spectrometry (LC/MS-MS) using validated analytical methods developed by MPI Research. Bioanalytics was conducted on samples from 4 animals per group (2 extra animals went through surgery and treatment to ensure that at least 4 per group would remain patent and without major tolerability issues).

## Results

**Table 1:**
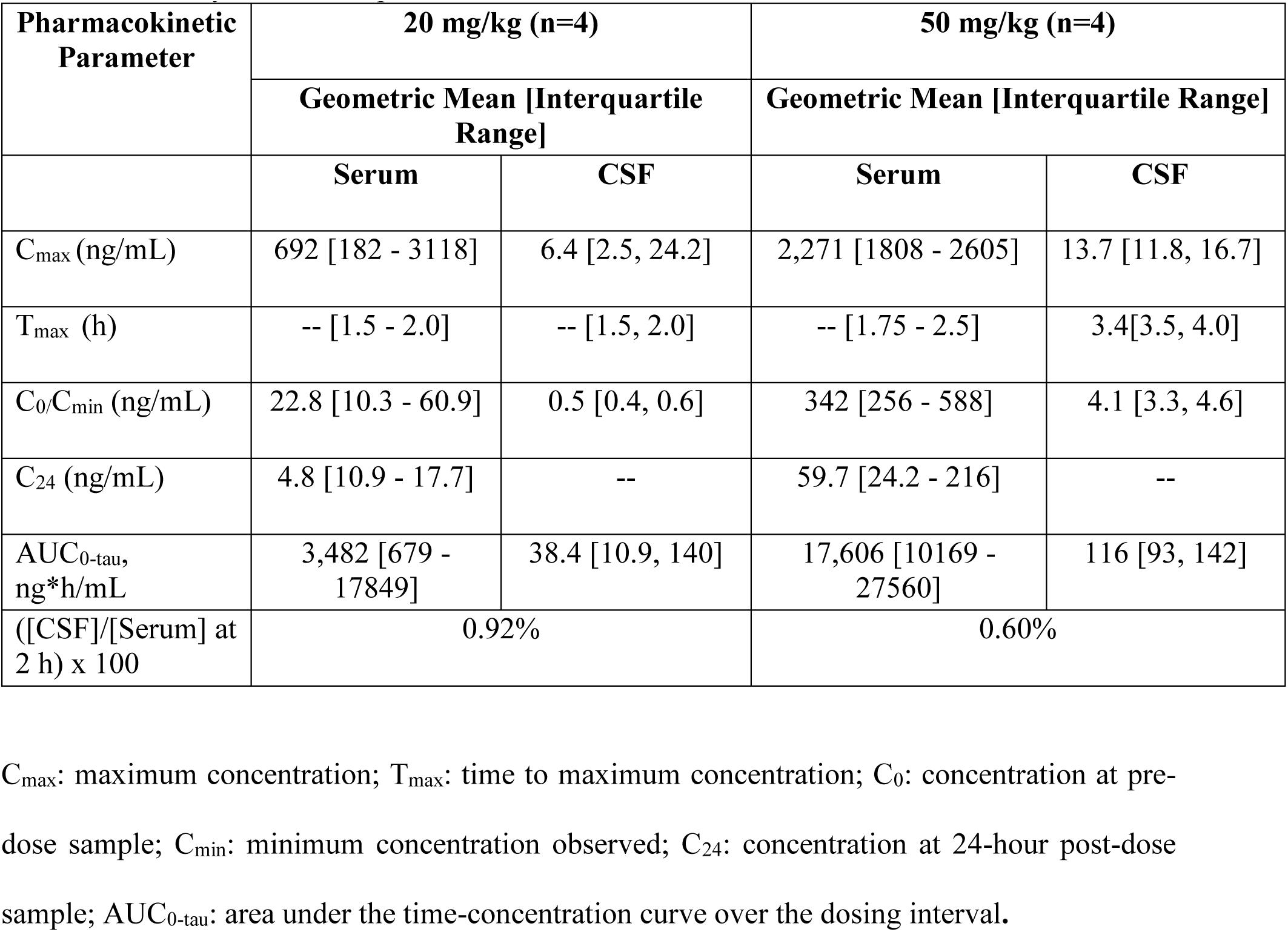
Summary of nilotinib pharmacokinetics in the serum and CSF.

High variability was seen in nilotinib concentrations in the 20 mg/kg group for both matrices for reasons unknown. The C_max_ concentration of nilotinib in the serum at 20 mg/kg was in the range of that observed in the 300 mg arm of the clinical study. As seen in the clinical data, the CSF levels of nilotinib were <1% of those observed in the serum.

### Brain Tissue Concentrations of Nilotinib in the Dog

On day 16, paired samples of serum and brain tissue were collected at 2 h post-dose to target the T_max_ concentrations. The average (standard deviations) serum concentrations in the 20 mg/kg and 50 mg/kg dose groups were 1359 (1318) ng/mL and 1214 (597) ng/mL, respectively. The average (standard deviations) brain tissue concentrations in the 20 mg/kg and 50 mg/kg dose groups were 4132 (4158) ng/g and 3355 (1392) ng/g, respectively. When corrected for protein binding (reported dog plasma protein binding is 98.3%; Xia et al., 2012), the mean (standard deviation) unbound concentration of nilotinib in the brain tissues were 70 (71) and 57 (24) ng/g tissue, which reflects 4% and 2.3% of serum concentration for 20 mg and 50 mg/kg arms, respectively.

**Assessment of c-Abl inhibition in the dog brain**.

## Methods

The dog brain tissues (cerebral cortex and cerebellum) were homogenized in RIPA buffer (50 mM Tris, pH 8.0, 150mM NaCl, 1% NonidetTM P-40, 1% SDS, 0.5% sodium deoxycholate) supplemented with phosphatase inhibitor cocktail II and III (Sigma-Aldrich), and complete protease inhibitor mixture. The homogenate was centrifuged (20 min at 4° C, 15000 rpm) and the resulting supernatant was collected. The protein concentrations of the samples were measured by BCA assay. Samples were electrophoresed on SDS-PAGE gels and transformed to nitrocellulose membranes. Membranes were blocked with 5% non-fat dry milk (wt/vol) in Tris-buffered saline with Tween-20 (TBS-T) and incubated with primary antibodies (mouse anti-c-Abl (# 554148, BD Biosciences); Rabbit anti-pY245 c-Abl (# 2861, Cell Signaling)). After an incubation with horseradish peroxidase-conjugated secondary antibody (anti-mouse IgG (# 7076S, Cell Signaling) or Anti-rabbit (# 7074S Cell Signaling)), the immunoblot signal was detected using chemiluminescent substrates (Thermo Scientific). The integrated band densities were measured using ImageJ software and the relative densities of pY245 c-Abl and c-Abl were calculated with respect to total c-Abl and beta-actin respectively.

## Results

Nilotinib treatment did not affect total c-Abl or pY245 c-Abl levels normalized to total c-Abl in the cortex or the cerebellum. The mean and (standard deviation) for the ratio of optical density of pY245 c-Abl to total c-Abl bands in the cerebral cortex for the vehicle, 20 mg/kg nilotinib and 50 mg/kg nilotinib groups, respectively, were: 2.133±0.82, 2.551±1.153 and 4.060±1.393. The corresponding values for the cerebellum were: 0.290±0.276, 0.766±0.544 and 0.792±0.444. Statistical analyses using Sidak’s multiple comparison test showed p>0.15 for the cortex and p>0.46 for the cerebellum.

Taken together, this carefully designed dog PK study with pharmacodynamic assessment indicates that nilotinib has poor brain penetration resulting in concentrations in the CNS that are not sufficient to inhibit c-Abl activity in the brain tissue.

## Other information

### Reproducible Research Statement

The following documents are available to the readers

Protocol: available at

Statistical Analysis Plan: available at

Statistical Code: available at

Data: will be uploaded on the NINDS clinical trials data repository

